# Risk factors for immune checkpoint inhibitor colitis: a retrospective multi-center cohort study using electronic health records

**DOI:** 10.1101/2025.11.05.25339621

**Authors:** Danny Hoi Tsun Chu, Ann-Kathrin Schalkamp, Vivek A. Rudrapatna

## Abstract

**Background:** Immune checkpoint inhibitors (ICIs) are effective for many cancers but often cause immune-related adverse events, particularly gastrointestinal (GI) complications such as colitis. Identifying risk factors for ICI colitis (CIC) could improve patient selection and discover potential mechanisms relevant to idiopathic inflammatory bowel disease (IBD). Prior studies on ICI risk factors have methodological limitations.

**Methods:** We used electronic health record data from six University of California institutions with 10,260 adults on ICIs. Baseline and time-varying predictors were defined based on prior IBD epidemiology studies, including interactions. LASSO Cox regression was applied to data from UCSF Central Data Warehouse to select the most predictive model, with coefficients estimated using multi-center data.

**Results and conclusion:** Fourteen significant predictors of CIC were identified. The strongest predictors were concurrent use of antibiotics (HR 2.72, 95% CI: 2.41–3.07) and non-steroid anti-inflammatory drugs (1.50, 1.16–1.94). Notable GI risk factors included gastroesophageal reflux disorder (1.31, 1.14–1.50), other GI disorders (1.32, 1.16–1.50), and disorders of the gallbladder, biliary tract, and pancreas (1.33, 1.15–1.57). High BMI before ICI administration increased risk (per unit 1.07, 1.04–1.09), while higher BMI after ICI administration decreased risk (per unit 0.93, 0.90–0.95). Melanoma, anxiety, and depression also increased risk. GI oncologists should consider ICI colitis in patients with comorbid GI disorders or those using antibiotics or NSAIDs. Baseline fecal calprotectin testing is recommended for these patients. Future studies are needed to explore the underlying mechanisms. Our findings suggest microbiome-and immune system–altering factors play roles in CIC and idiopathic GI disorders.

## INTRODUCTION

Immune checkpoint inhibitors (ICIs) can significantly improve survival outcomes for cancer patients, however, their use is limited by serious immune-related adverse events (irAEs) occurring in roughly 40% of patients [1]. By targeting key immune regulatory pathways, such as programmed death-1 (PD-1) and cytotoxic T lymphocyte antigen-4 (CTLA-4), ICIs enable the immune system to recognize and attack tumor cells more effectively [2] and are commonly used for melanoma, renal cell carcinoma, head and neck cancers and non-small cell lung cancer [3]. Despite their effectiveness, in general, irAE occurred in 40.0% (95% CI: 37.3 – 42.7%) of patients, with 12.5% being gastrointestinal related irAEs (IQR: 5.7 – 33.3%) [1]. ICI-induced colitis (CIC) is the most reported irAE. Beyond reducing the already diminished quality of life of these patients, CIC can result in hospitalization, the need for additional immunosuppressive treatments, surgery, and even death [4–6]. Understanding risk factors that predispose patients to CIC can improve patient selection, inform early treatment and risk-mitigation strategies and ultimately improve patient outcomes. Moreover, a greater understanding of disease risk factors can provide insights into the pathophysiology of CIC itself as well as related disorders like idiopathic inflammatory bowel diseases (IBD)[7].

Previous research has identified several clinical risk factors for ICI colitis. These include the choice of ICI (anti-CTLA4 as more risky than anti-PD1/L1) and use of combination treatment, female sex, younger age, non-steroid anti-inflammatory drugs (NSAIDs) use, and antibiotics use [8–16]. While these studies have contributed to our understanding of ICI pathophysiology, their interpretation is somewhat limited by their use of single-center data and homogeneous populations with uncertain generalizability. Moreover, prior studies have not utilized longitudinal nature modeling techniques to account for the sequential nature of clinical decision making after starting ICIs. Longitudinal models like time-dependent cox regression can help reduce length-time bias and optimally estimate the total effect of time-varying exposures on the risk of developing colitis.

Here, we used real-world EHR to investigate risk factors of developing colitis for cancer patients receiving ICI. We consider a wide range of potential risk factors informed by prior research on inflammatory bowel disease. We performed feature selection with the data from University of California, San Francisco (UCSF) Central Data Warehouse (CDW) and present the results of the Cox regression in a cohort encompassing six institutions extracted from the University of California Data Discovery Platform (UCDDP).

## METHODS

This study utilized deidentified electronic health records data from the University of California Health Data Warehouse, and thus was deemed suitable for expedited review by the UCSF IRB.

### Cohorts

Our cohort criteria were the same for the UCSF and the UCDDP cohorts between January 2012 and March 2025. Inclusion criteria were as follows: 1) diagnosis of either skin cancer, lung cancer, renal cell carcinoma, or head and neck cancer with at least one administration of any ICIs, 2) older than 18 years at the first administration of ICI. The date of first ICI administration forms the index date. The exclusion criteria were: 1) patients with a history of inflammatory bowel disease, 2) patients with a history of GI-related tumors, and 3) any pregnancy or abortion during the study period.

### Predictors and Outcome Variables

Information on demographics, medication history, diagnosis history, and details on ICI administration were extracted from the structured EHR data. We began with 44 candidate predictors and defined 7 interaction terms (Table 1). 21 predictors were identified as time-varying. We used ICD-9, ICD-10, and regular expressions to extract diagnoses-related predictors (Supplemental Table 1).

**Table 1.** Predictors included for feature selection. We list all predictors considered for feature selection and indicate their encoding (binary, categorical, continuous). * Refer to Supplemental Table 1 for the definition in ICD code ^¶^ Anti-PD-1, Anti-PD-L1 or Anti-CTLA-4. Interaction between two agents if combination therapy was administered. The interaction of sex with combination therapy was noted as a three-variable interaction. The accumulated number of administration for combination therapy was separated out, but the component of the regimen was recorded into accumulated dose of anti-PD1/L1 and anti-CTLA-4 as well. Moreover, due to long half-life of ICIs, their presentation in the dataset will be carried forward to the following month even if there is no administration in the following month[17].

| Demographics | Health history/conditions | ICI administration | Medication / procedure history | Interaction |
| --- | --- | --- | --- | --- |
| Sex- Female (Binary)<br>Race: White (Binary)<br>Age (Continuous)<br>Smoking Status (Categorical)<br>Driving Miles to San Francisco (SF) zip 94110 (Continuous)<br>Driving hours to SF zip 94110 (Continuous)<br>Area Deprivation Index State Rank in California (Ordinal)<br>Marital Status (Categorical) | BMI (time-varying) (Continuous)<br>BMI (before initiation of ICI) (Continuous)<br>Immune disorder (Binary)*<br>Gastroesophageal reflux disorder (Binary)*<br>Gastrointestinal ulcer (Binary)*<br>Liver disease (Binary)*<br>Disorder of gallbladder, other biliary tract, or pancreas (Binary)*<br>Other gastrointestinal disorder (Binary)*<br>Melanoma (Binary)*<br>Other skin tumour (Binary)*<br>Non-small cell lung cancer (Binary)*<br>Small cell lung cancer (Binary)*<br>Renal Cell Carcinoma (Binary)*<br>Head and neck cancer (Binary)*<br>Appendectomy (Binary)<br>Depression (Binary)*<br>Bipolar disorder (Binary)*<br>Anxiety (Binary)*<br>Schizophrenia and related psychosis (Binary)*<br>Other mood disorder (Binary)* | Type of ICI administered (Categorical) <sup>¶</sup><br>Accumulated number of administrations for each type of ICI (Ordinal) <sup>¶</sup> | Antibiotics administration (Binary)<br>Non-steroid anti-inflammatory drug administration (Binary) | Female x Type of ICI administered (Binary) <sup>¶</sup><br>Female x Accumulated number of administrations of each type of ICI (Ordinal) <sup>¶</sup> |

The outcome variable was defined as any non-infective colitis that occurred after administration of ICI, defined by ICD-10 code of K50, K51 or K52. Competing events were defined as death, treatment discontinuation, and non-GI-related adverse events leading to discontinuation. Treatment failure was defined as having a record of other anti-neoplastic drugs after the last recorded ICI dose. Non-GI-related adverse events were defined as any rash, hypothyroidism, pneumonitis, nephritis, hypophysitis, adrenal insufficiency, type 1 diabetes, myalgia, or encephalopathy alongside no further administration of ICI.

### Data cleaning and imputation

Both the UCSF CDW and UCDDP datasets were condensed to monthly records. If multiple visits occurred during one calendar month, more recent information was prioritized and missingness was filled with prior records of that month. Overall, missingness of the identified predictors was low (less than 3%) (Supplemental Table 2 and 4).

Due to the discrete representation of time and the inclusion of time-varying predictors, it could occur that both a new record and the end of the observation fell into the same month, leading to tstart=tend. To address this, we used a split-second offset of 0.001 to ensure that all time intervals were greater than zero.

Last observation carried forward was applied for the accumulated number of administrations of ICIs and all the diagnoses. If no record was found for medication or diagnosis, they were assumed to be absent, recorded as “0”.

Apart from the time-varying BMI, which was imputed by person-wise linear interpolation with previous and future data, the rest of the covariates were imputed using random forest as implemented in the R package `mice`. Missing data were imputed in two stages to appropriately handle both baseline and time-varying covariates. For non–time-varying covariates, imputation was based solely on baseline records (i.e., the first observation for each participant) to ensure consistent values across all time points and to avoid biasing the imputation toward participants with more frequent records. In contrast, time-varying covariates were imputed using the entire dataset to capture temporal variability and population-wide patterns. Imputation was done 5 times with 10 iterations each. Numeric data was incorporated by taking the mean of the imputations, ordinal data was incorporated by taking the median of the imputations, and categorical data was incorporated by taking the mode of the imputations.

### Statistical analysis

Using the UCDDP cohort, we computed the overall incidence rate and each institution’s. Confidence intervals were calculated assuming Poisson Distribution.

We used LASSO time-dependent Cox regression to perform feature selection in the UCSF CDW dataset. The selected feature set was formed by removing those with zero estimates and those with more than 10% missingness in the UCDDP dataset (for example, residential information and smoking).

The selected features were used in the UCDDP dataset in a time-dependent Cox regression to estimate the hazard ratio for each predictor. P-values were adjusted for multiple comparisons using the False Discovery Rate (FDR), with a significance threshold of α = 0.05. Only variables below this threshold will be reported, unless otherwise stated. The interaction terms were handled with R package of `biostat3` to produce a more interpretable sex-specific hazard ratio.

Further, a time-dependent Cox regression on competing events, including death and all-cause treatment discontinuation was fitted. In case any protective effect was shown in the main Cox regression for colitis, the same covariate was checked in the Cox regression on competing event to ensure the protective effect was not due to competing event.

Post-hoc analysis was performed to assess the differences between institutions. We trained an additional model, including institutions as dummy coded variables, and compared the AIC and BIC to the model without this information. We further evaluated the change in hazard ratios between the two models.

We further investigated the diseases captured under the umbrella predictor variable “other GI disorder” to indicate driving factors. For this, we identified 38 subgroups, each containing at least 10 patients, of diagnoses via ICD code hierarchy and calculated for each the frequency of colitis.

## RESULTS

### Feature selection

25 out of 46 features were selected through the LASSO time-dependent Cox regression (Supplemental Table 4, Supplemental Fig 3). Refer to Supplemental Fig 1, 2 and Supplemental Table 2, 3 for details on the UCSF cohort.

Features that were dropped by the LASSO time-dependent Cox regression included white race, female sex, smoking status, age, PD-1/L1, the accumulated number of anti-CTLA-4 administrations, the accumulated number of combined therapy administrations, ulcers, irritable bowel syndrome, liver disease, RCC, head and neck cancer, appendectomy, other mood disorder, as well as the interaction terms involving female x anti-PD-1/L1 x anti-CTLA-4, female x accumulated anti-PD-1/L1 administrations, female x accumulated anti-CTLA-4 administration, female x accumulated combined therapy administration, and anti-PD-1/L1 x anti-CTLA-4 (Supplemental Table 3).

Residential information, while being selected as contributing risk factors, was not available or had high missing rates in UCDDP and was thus excluded from further analysis. The UCSF model attributed low effect to these variables.

### Clinical characteristics

Across the six institutions in UC, we identified 20,413 patients who received checkpoint inhibitors, of which 12,295 had at least one of the included cancers as well as no history of GI-related tumors. We further excluded 20 patients who were younger than 18 years before their first administration of ICI, 1,251 with a previous history of non-infectious colitis, 745 with recorded pregnancy/abortion, and 14 with missing data about the timing of ICI administration or no record after ICI administration. Our final cohort included 10,265 patients (Fig. 1), 4,040 (39.4%) females, median age of 68.8 (IQR: 60.0 – 77.2), with a median follow-up of 7 months (range: <1-148 months) since first ICI administration (Table 2). 1,147 (11.2%) developed colitis during follow-up (Fig 2), and 2,765 (26.9%) experienced competing events such as death, other adverse events leading to treatment cessation, or treatment failure (see Methods). Refer to Supplemental Table 6 for details on missingness.

**Fig 1.**
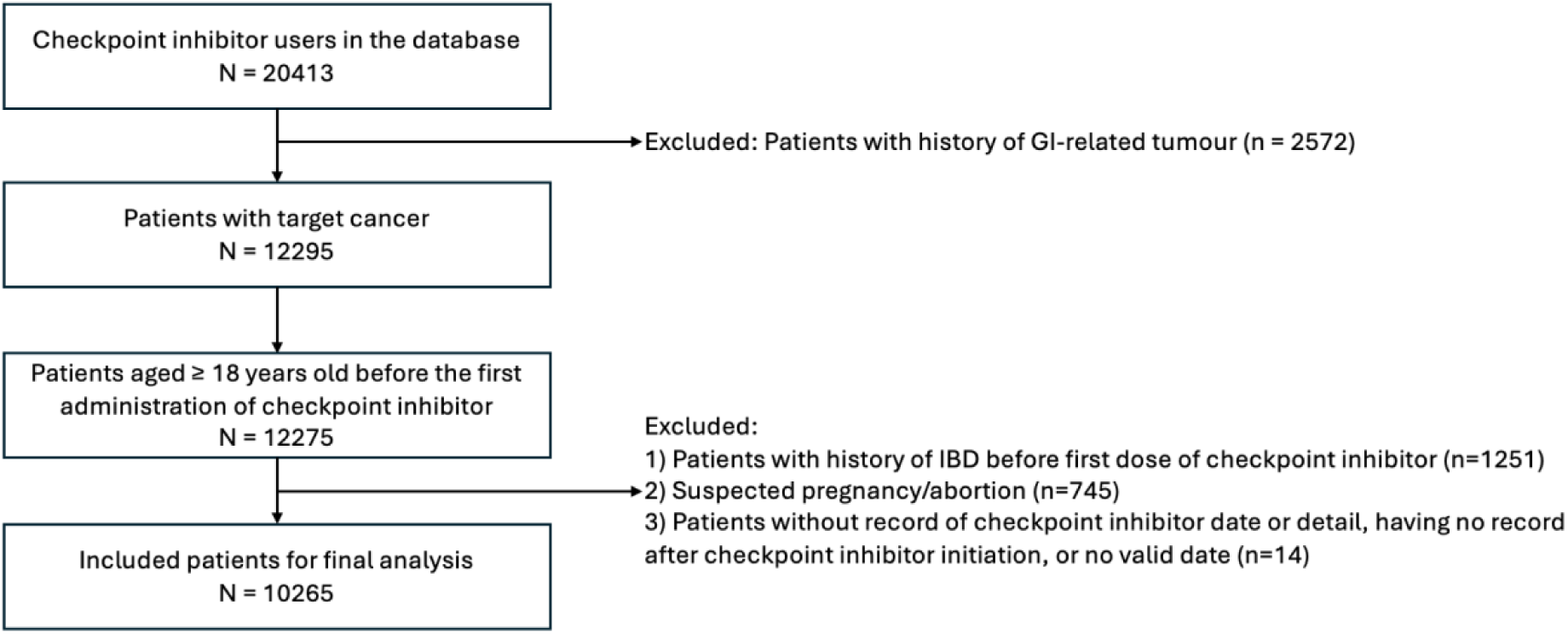
Flow chart of patient selection and data extraction For the UCDDP cohort we show the initial cohort size and the amount of individuals dropped due to inclusion and exclusion criteria.

**Fig 2.**
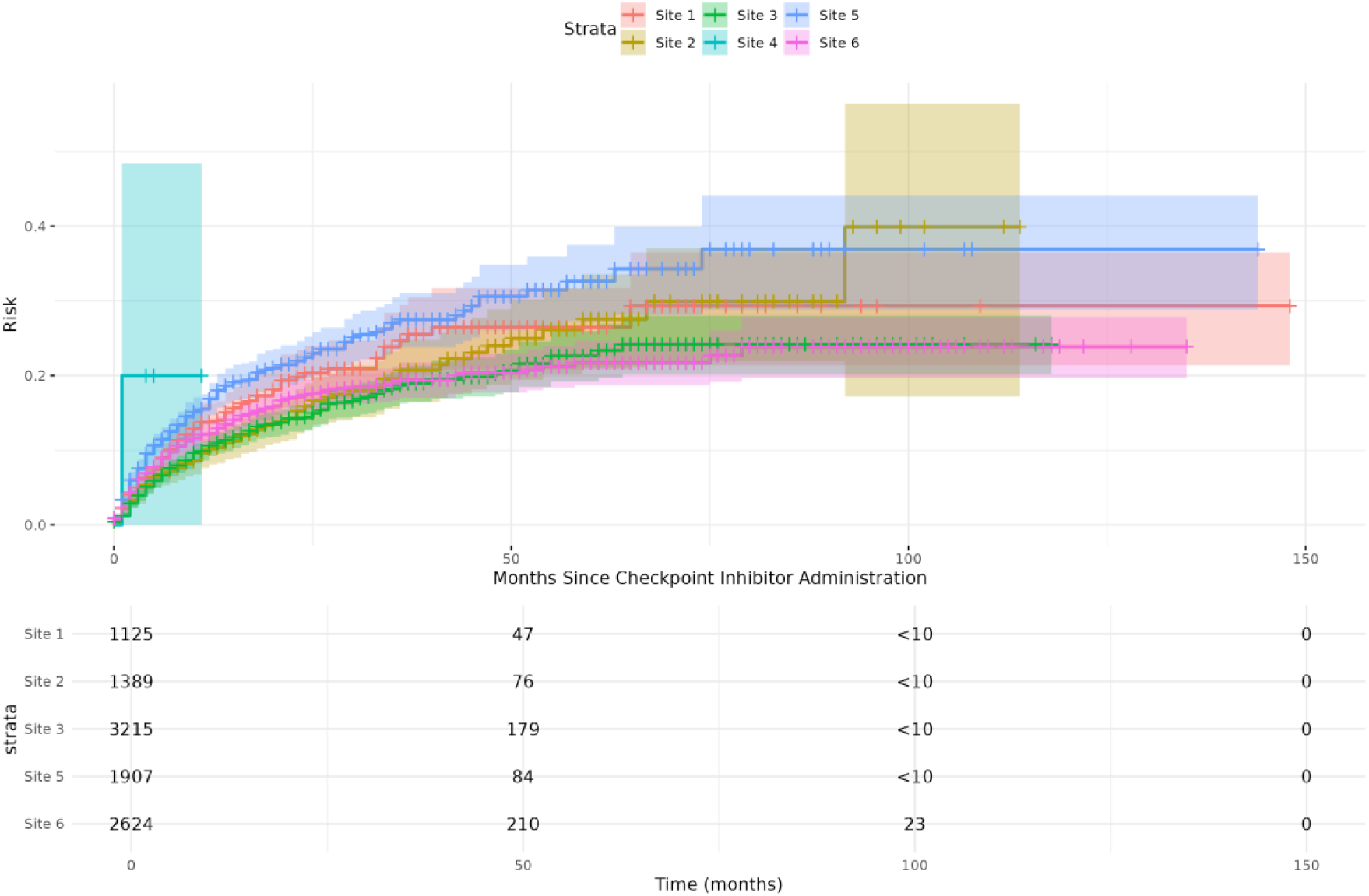
Cumulative incidence of colitis following checkpoint inhibitor administration by institution

**Fig 3.**
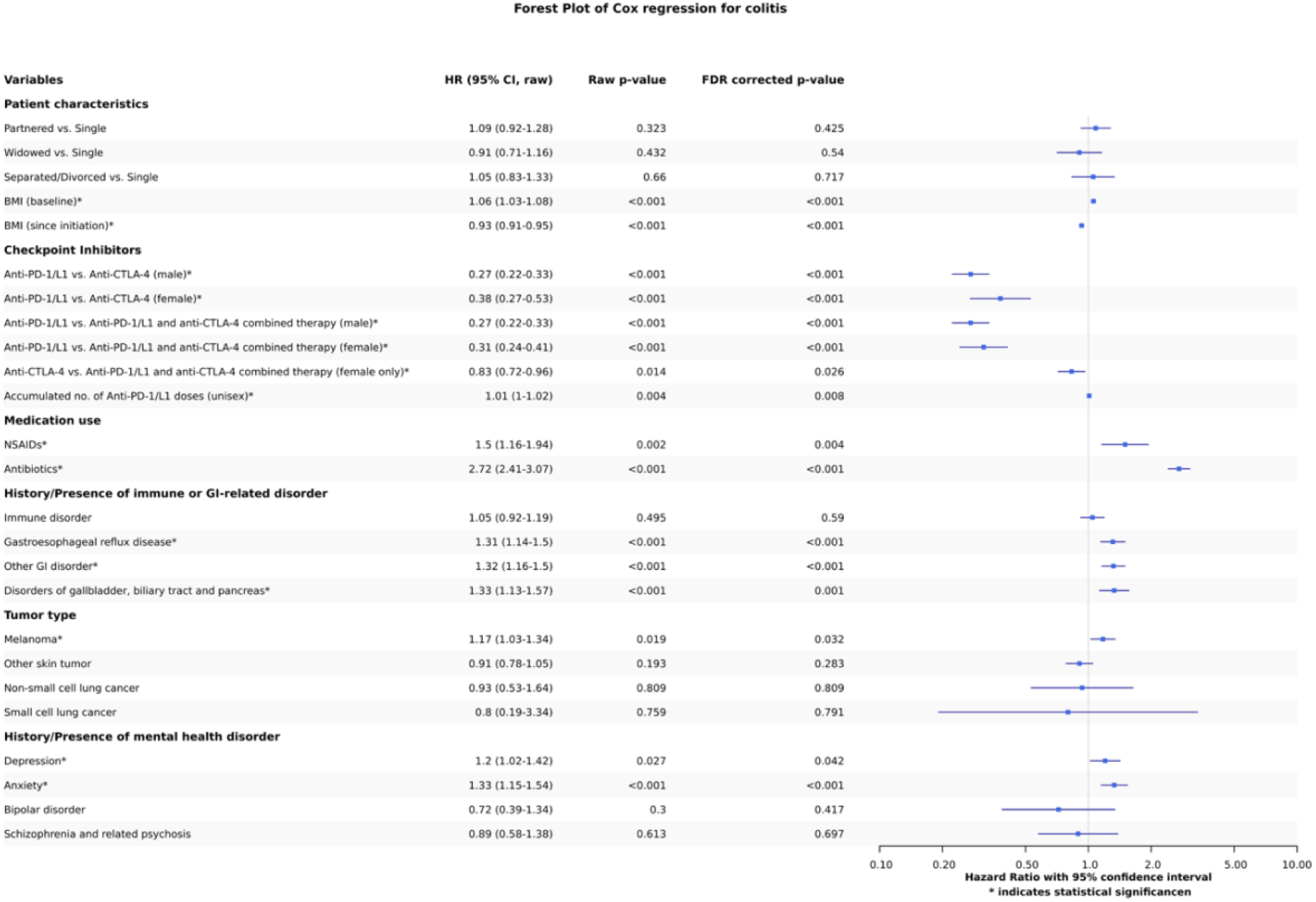
Forest plot of the time-dependent Cox regression model applied

**Table 2.**
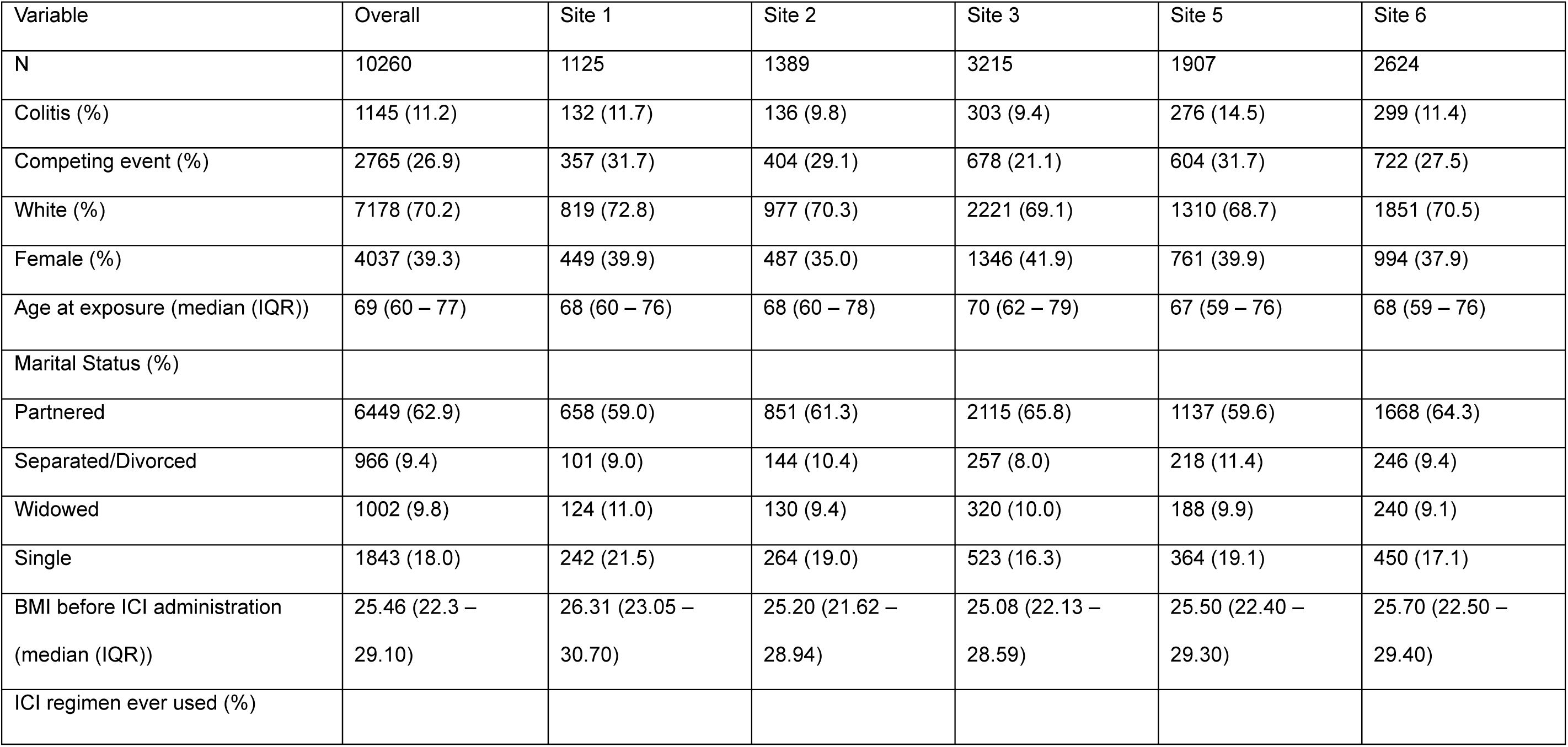
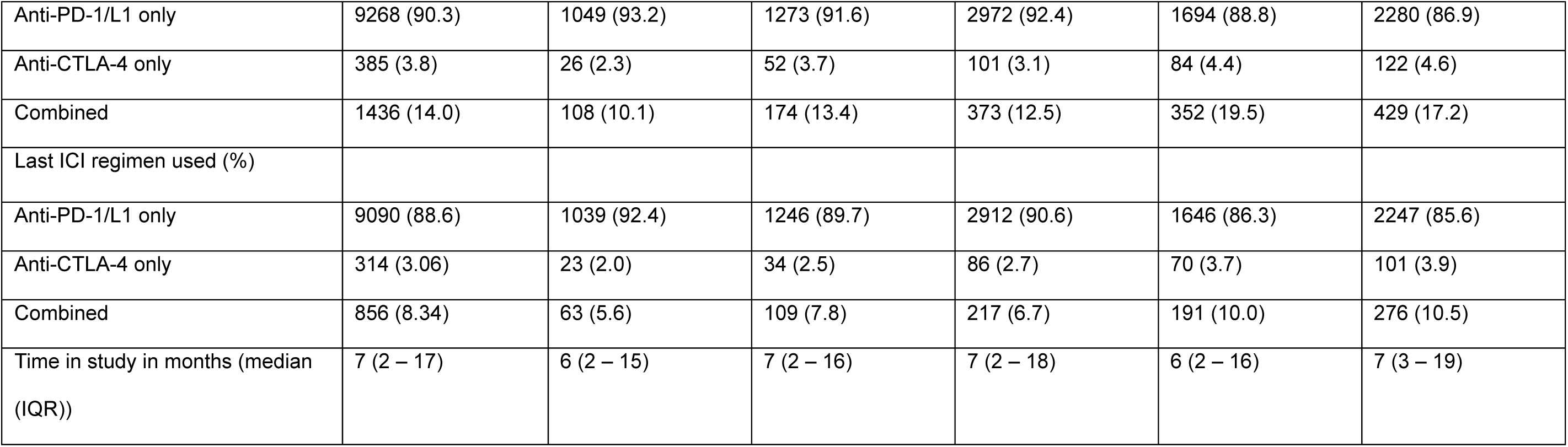
Cohort characteristics by institution. The demographics and characteristics of the identified UCDDP cohort are shown. The overall cohort is split by institution to demonstrate each institution’s demographics. Site 4 was removed from the table due to a patient count of less than 10. BMI: Body Mass Index; ICI: Immune checkpoint inhibitor; IQR: Inter-quartile range

| Variable | Overall | Site 1 | Site 2 | Site 3 | Site 5 | Site 6 |
| --- | --- | --- | --- | --- | --- | --- |
| N | 10260 | 1125 | 1389 | 3215 | 1907 | 2624 |
| Colitis (%) | 1145 (11.2) | 132 (11.7) | 136 (9.8) | 303 (9.4) | 276 (14.5) | 299 (11.4) |
| Competing event (%) | 2765 (26.9) | 357 (31.7) | 404 (29.1) | 678 (21.1) | 604 (31.7) | 722 (27.5) |
| White (%) | 7178 (70.2) | 819 (72.8) | 977 (70.3) | 2221 (69.1) | 1310 (68.7) | 1851 (70.5) |
| Female (%) | 4037 (39.3) | 449 (39.9) | 487 (35.0) | 1346 (41.9) | 761 (39.9) | 994 (37.9) |
| Age at exposure (median (IQR)) | 69 (60 – 77) | 68 (60 – 76) | 68 (60 – 78) | 70 (62 – 79) | 67 (59 – 76) | 68 (59 – 76) |
| Marital Status (%) |  |  |  |  |  |  |
| Partnered | 6449 (62.9) | 658 (59.0) | 851 (61.3) | 2115 (65.8) | 1137 (59.6) | 1668 (64.3) |
| Separated/Divorced | 966 (9.4) | 101 (9.0) | 144 (10.4) | 257 (8.0) | 218 (11.4) | 246 (9.4) |
| Widowed | 1002 (9.8) | 124 (11.0) | 130 (9.4) | 320 (10.0) | 188 (9.9) | 240 (9.1) |
| Single | 1843 (18.0) | 242 (21.5) | 264 (19.0) | 523 (16.3) | 364 (19.1) | 450 (17.1) |
| BMI before ICI administration<br>(median (IQR)) | 25.46 (22.3 –<br>29.10) | 26.31 (23.05 –<br>30.70) | 25.20 (21.62 –<br>28.94) | 25.08 (22.13 –<br>28.59) | 25.50 (22.40 –<br>29.30) | 25.70 (22.50 –<br>29.40) |
| ICI regimen ever used (%) |  |  |  |  |  |  |
| Anti-PD-1/L1 only | 9268 (90.3) | 1049 (93.2) | 1273 (91.6) | 2972 (92.4) | 1694 (88.8) | 2280 (86.9) |
| Anti-CTLA-4 only | 385 (3.8) | 26 (2.3) | 52 (3.7) | 101 (3.1) | 84 (4.4) | 122 (4.6) |
| Combined | 1436 (14.0) | 108 (10.1) | 174 (13.4) | 373 (12.5) | 352 (19.5) | 429 (17.2) |
| Last ICI regimen used (%) |  |  |  |  |  |  |
| Anti-PD-1/L1 only | 9090 (88.6) | 1039 (92.4) | 1246 (89.7) | 2912 (90.6) | 1646 (86.3) | 2247 (85.6) |
| Anti-CTLA-4 only | 314 (3.06) | 23 (2.0) | 34 (2.5) | 86 (2.7) | 70 (3.7) | 101 (3.9) |
| Combined | 856 (8.34) | 63 (5.6) | 109 (7.8) | 217 (6.7) | 191 (10.0) | 276 (10.5) |
| Time in study in months (median (IQR)) | 7 (2 – 17) | 6 (2 – 15) | 7 (2 – 16) | 7 (2 – 18) | 6 (2 – 16) | 7 (3 – 19) |

**Table 3.**
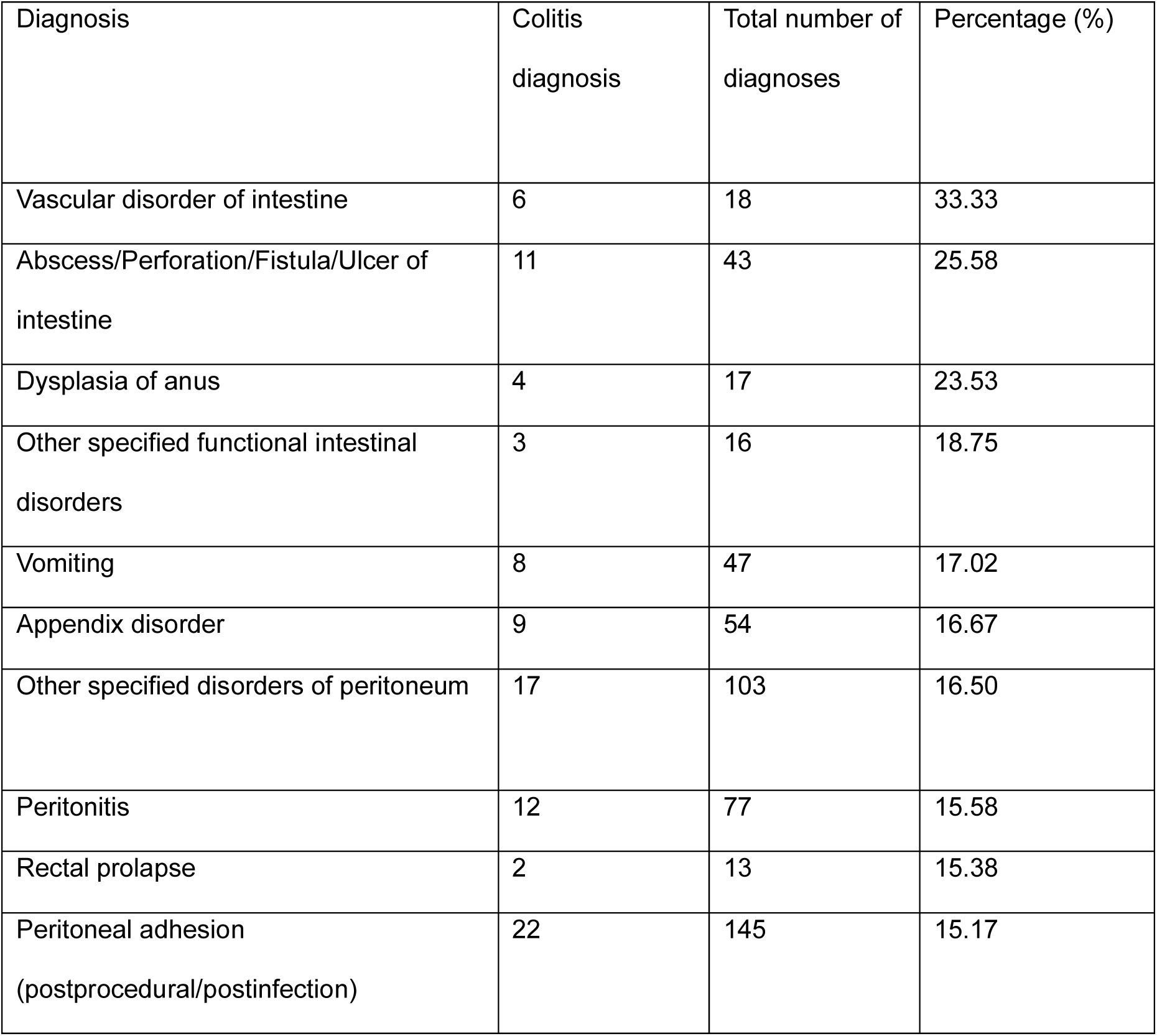
Colitis diagnosis proportion for each of the top 10 diagnosis in other GI disorder. The table listed the top 10 rate of colitis diagnosis in the sub-cohort of people diagnosed with GI disorder. The p-value was calculated compared to the rate of colitis in all of the people with other GI disorder

| Diagnosis | Colitis diagnosis | Total number of diagnoses | Percentage (%) |
| --- | --- | --- | --- |
| Vascular disorder of intestine | 6 | 18 | 33.33 |
| Abscess/Perforation/Fistula/Ulcer of intestine | 11 | 43 | 25.58 |
| Dysplasia of anus | 4 | 17 | 23.53 |
| Other specified functional intestinal disorders | 3 | 16 | 18.75 |
| Vomiting | 8 | 47 | 17.02 |
| Appendix disorder | 9 | 54 | 16.67 |
| Other specified disorders of peritoneum | 17 | 103 | 16.50 |
| Peritonitis | 12 | 77 | 15.58 |
| Rectal prolapse | 2 | 13 | 15.38 |
| Peritoneal adhesion (postprocedural/postinfection) | 22 | 145 | 15.17 |

The plot shows the cumulative incidence of colitis since first ICI administrations for each UC hospital separately. Site 4 (<10 patients) and other time points with less than 10 patients were not shown in the risk table due to risk of identification.

### Incidence Rate of Colitis

The identified UC-wide incidence rate of colitis after the first ICI administration was 9.54 episodes (95% CI: 9.21 – 10.40) per 100 person-year. The institutions showed varying, but overlapping, incidence rates ranging from Site 3 with 8.10 (95% CI: 7.21 – 9.07) to Site 4 with 46.2 (95% CI: 1.17 – 257.15) per 100 person-year (Supplemental Table 5). The median time to colitis onset was 5 months (IQR: 2 - 11 months).

### Identified risk factors for colitis

23 predictors were included in the time-dependent Cox regression and their effect on colitis risk was assessed. 13 of those were statistically significant after FDR correction, including 1 interaction terms.

The largest hazard ratio was identified for the binary indicator of the concurrent use of antibiotics (HR: 2.72, 95% CI: 2.41– 3.07). In terms of patient characteristics, BMI for both baseline (HR: 1.06 (1.03 – 1.08)) and time-varying after ICI initiation (HR: 0.93 (0.91 – 0.95)) indicated a statistically significant difference. For the choice of the ICI agent, anti-PD-L1 had a generally lower risk of colitis when compared to anti-CTLA-4 (Male HR: 0.27 (0.22 – 0.33); Female HR: 0.38 (0.27 - 0.53)) and combined therapy (Male HR: 0.27 (0.22 – 0.33); Female HR: 0.31 (0.24 - 0.41)). For females, it was found that anti-CTLA-4 had a lower risk of colitis than combination therapy as well (Female HR: 0.83 (0.72 - 0.96)). The interaction between female and anti-PD-1/L1 choice was also detected as a statistically significant difference (HR: 1.20 (1.04 – 1.39)).

The accumulated number of doses of both anti-PD-1/L1 (HR: 1.01 (1.00 – 1.02)) was identified as a significant risk factor. Medication usage apart from antibiotics, NSAIDs use was found to increase colitis risk (HR: 1.5 (1.16 – 1.94)). The presence of GERD (HR: 1.31 (1.14 – 1.50)), other GI disorder (HR: 1.32 (1.16 – 1.50)), and disorders of gallbladder, biliary tract and pancreas (HR: 1.33 (1.13 – 1.57)) increased risk of colitis. Further, having melanoma as the type of cancer (HR: 1.17, (1.03 – 1.34)) and having depression (HR: 1.20 (1.02 – 1.42)) or anxiety (HR: 1.33 (1.15 – 1.54)) significantly increased the risk of colitis.

The estimated coefficients of the time-dependent Cox regression model are shown with their hazard ratio (HR) and respective 95% Confidence interval as well as their uncorrected and FDR-corrected p-value. The x-axis is presented on a log scale.

### Post-hoc analysis: Assessing the difference between institutions

As the UCDDP cohort encompasses six different institutions, we assessed the influence of the institution. Comparing the estimated survival rates across institutions with log rank test showed statistically significant differences (p<0.001). We thus added institution as a covariate in a separate Cox regression, which resulted in an increase in the AIC by 15 (from 18664.96 to 18850.40), and an increase in BIC by 11 (from 18980.93 to 18991.58). None of the institutions contributed significantly to the colitis risk with the estimated hazard ratios for the institutions compared to Site 6 being Site 1: 1.11 (95% CI: 0.91 – 1.37, p = 0.779), Site 2: 0.81 (0.66 – 1.00; p = 0.085), Site 3: 0.85 (0.72 – 0.99; p = 0.084), Site 4: 4.32 (0.60 – 39.93, p = 0.225) and Site 5: 1.18 (1.00 – 1.41; p = 0.085). The hazard ratio changes of the other predictors in percentage ranged from-3.03% to 2.66% compared to the previous results (Supplemental Table 7).

### Post-hoc analysis: Assessing other GI disorder by diagnosis

With other GI disorders being selected as an important risk factor but representing various diseases, we further identified the diagnoses included in this umbrella term with the highest colitis rates. In total, 673 people out of 4,521 people (14.89%) diagnosed with “other GI disorder” developed colitis.

Vascular disorder of intestine had the highest colitis rate (6/18, 33.33%), followed with Abscess/Perforation/Fistula/Ulcer of intestine (11/43, 25.58%) and dysplasia of anus (4/17, 23.53%). The full table can be found in Supplemental Table 8.

## DISCUSSION

In this large UC-wide cohort study, we explored various predictors for their association with colitis risk following ICI administration. Our analysis verified previously identified risk factors but further highlighted additional variables to consider for patient selection and monitoring during administration of ICI.

Our analysis reproduced several previous findings, conferring robustness to those findings through the use of a large and demographically diverse cohort. In particular, we verified that anti-PD-1/L1 therapy was associated with a lower risk of immune-related colitis compared to anti-CTLA-4 monotherapy, a pattern consistent across both male and female patients [18–24]. An increased risk for the melanoma cancer type was also reproduced [25–27]. Moreover, baseline obesity or overweight patients having a higher risk of irAEs or colitis was also reproduced in our cohort with the baseline numeric BMI being selected as a risk factor[28, 29]. We have also reproduced the positive association between depression, anxiety and colitis, which agrees with previous findings on psychological risk factors [30, 31]. Pancreatic, gallbladder, and biliary tract disease has not been extensively studied as a risk factor for irAE or colitis but has been mentioned as a risk factor for IBD[32]. Our study identified them as risk factors for ICI colitis.

BMI changes during ICI treatment was identified as a risk factor for colitis in our study without increasing the risk of competing risk (Supplemental Fig 4). While previous research has investigated baseline BMI or obesity, our study investigated the dynamics of BMI. This weight loss could be explained by reverse causation meaning that symptoms of colitis can cause weight loss, for instance through diarrhea, loss of appetite, and nausea [5, 14]. However, prior research suggests that after minimizing the effect of reverse causation with IBD, weight gain since early adulthood may be associated with a lower risk of microscopic colitis [33]. Our results suggest that BMI should be monitored in patients administered with ICI, and additional testing for colitis should be carried out for patients with a BMI drop. For instance, any symptoms, such as diarrhea and abdominal pain, should be further investigated with endoscopy, histology or biopsy[34].

Sex-specific differences emerged when comparing combination therapy with anti-CTLA-4 monotherapy. Among female patients, anti-CTLA-4 alone was associated with a lower risk of colitis than combination therapy, in addition to the number of administrations of anti-PD-1/L1. Although there were no literature investigated the interaction of each agent with sex, the trend of our study supported what was suggested male has a lower risk of colitis than female in terms of ICI irAEs that includes GI via the interaction [35, 36]. Thus, we suggest that sex-specific interaction with choice of ICI type should be further studied.

Our study also found an increasing risk of colitis with the number of administrations of anti-PD-1, regardless of whether it is in monotherapy or combined therapy. Although other studies found conflicting results with dose or total infusions, using Cox regression tackled the issue of length-time bias as the accumulated number of administrations was treated as a time-varying covariate[18, 37]. However, given that there is only a weak association, the difference will be less apparent per dose, and this may not be as critical as other predictors while allocating resources to monitor. With the result of accumulated number of administrations for anti-CTLA-4 or combined therapy being selected out during LASSO, we suggest early and close monitoring for any regimen containing anti-CTLA-4 compared to anti-PD1/L1 monotherapy, regardless of the number of administrations.

Antibiotic use and NSAIDs use were also previously reported for either irAEs in general or colitis specifically [14, 38]. In our study, these had the largest hazard ratios. Interestingly, previous studies identified a lower occurrence and recurrence rate of ICI for antibiotics used at any time (including the time before ICI initation and after ICI discontinuation), and an increased risk of severe colitis, especially with those targeting anerobic activity, which agrees with our findings [39]. As we incorporated antibiotics and NSAIDs use as time-varying predictors that precisely indicate timepoints of concurrent usage with ICI, our findings indicate a risk of concurrent usage. It is unclear from our results whether the usage of antibiotics or the infection being treated is increasing the risk for colitis. Further research is required to disentangle this.

Novel predictors identified in our analysis were the binary indicator for the presence of GERD, “other GI disorders”, and “disorders of gallbladder, biliary tract and pancreas”. The influence of prior GI disorders (except IBD) on colitis has not been studied extensively. Our findings suggest that, in general, pre-disposing GI disorders increased the risk for CIC. Those prior GI disorders could be an indicator of underlying gastrointestinal vulnerability with ICI triggering colitis development [40, 41]. Previous studies have investigated histopathology for identifying colitis that will improve once ICI treatment is stopped versus colitis that will prevail even after ICI treatment cessation. They identified basal lymphoplasmacytosis [41, 42] to differentiate different IBD on whether it is self-limiting, chronic IBD triggered by ICI, or idiopathic IBD. The type of colitis that does not stop once treatment is stopped was referred to as underlying-IBD, behaving similarly to chronic IBD and thus supporting this notion of the ICI treatment triggering the IBD in some individuals. Our findings support this hypothesis of ICI acting as a trigger in those pre-disposed people, and thus, anyone with a previous GI disorder should have extra monitoring for the risk of colitis development while being administered ICI. Post-hoc analysis showed that the frequency of colitis varied from 3.45% to 33.33% among the diseases included under the’other GI disorder‘umbrella term. It was found that those groups with the highest rates were related to underlying mucosal damage in the lower GI tract. A previous study suggested a shorter time-to-colitis for patients with pre-existing IBD [43]. We could not verify this as we excluded patients with prior IBD diagnosis from our cohort due to it being indifferentiable from colitis in our database. In general, further research into the pathophysiology of colitis and IBD is needed, with our prior studies suggesting a hypothesis of underlying disease triggered through ICI or other disruptors. Ensuring generalizability across hospital settings and populations is crucial when incorporating risk factors into clinical practice. Our post-hoc analysis across the included six different institutions identified no significant difference between institutions. This suggests that our reported findings are robust across the six included institutions and likely generalize well to other cohorts. While previous studies were usually done in single-centre settings, there were only a few studies that were done across multiple centers in various healthcare institutions with a large sample size[13]. Our analysis, while being limited to UC hospitals, provides a large sample size (N=10,260) with a diverse population.

Our study has several limitations that have to be considered when interpreting results. While we included data from six institutions with the aim of covering a goodrepresentation of the global population, our data was limited to the UC system, which is geographically bound to California, so the generalizability to other states or countries cannot be guaranteed.

While we considered a considerable number of predictors, several variables could not be included in the analysis due to high rates of missingness, such as vitamin D level and cytokines, as well as metastasis status and cancer stage. We used a 1-month interval to define clinically significant ICI retention, with later colitis diagnoses after ICI cessation not being considered in our analysis. Given the 2 to 4-week half-life and the pharmacological principle that five half-lives are required for complete drug clearance, ICI exposure may remain clinically significant for longer than one month [17]. Our outcome variable to define colitis was based on a more broad definition of colitis, including any IBD diagnosed after initiation of ICI. We did not solely use the reporting of toxic colitis to determine the outcome due to varying reporting standards and this information being captured in many multiple fashions. As we relied on accurate date of diagnosis and ICI administration information in the EHR to determine prior or incident colitis, we potentially include some individuals in our dataset with colitis prior to ICI administration whose diagnosis was not accurately captured in the EHR data. Our analysis and results rely on the accuracy of the data available in EHR.

## Conclusion

To our knowledge, this is the first study examining time-to-colitis in a large, multi-center cohort with Cox models and numerous predictors. Our findings replicated known risk factors such as white race and baseline BMI, but also highlighted that pre-existing GI disorders, decrease in BMI, concurrent NSAID and/or antibiotics use to be significant risk factors for colitis. Those variables should be considered for treatment selection and monitored during ICI administration to reduce colitis risk and improve patient outcomes.

## Supporting information

All supplemental files

## Data Availability

Access to UCDDP and UCSF CDW database are controlled due to sensitive nature of the patient data, and they were protected by law. The UCSF CDW database can be accessed by UCSF-affiliated individuals by contacting UCSF Clinical and Translational Science Insitute or UCSF‘s Information Commons team, more information could be found at https://data.ucsf.edu/research/ucsf-data. If the reader is unaffiliated with UCSF, they can set up an official collaboration with a UCSF-affiliated investigator by contacting the principal investigator. UCDDP is only available to UC researchers who have completed analyses in their respective UC first and have provided justification for scaling their analyses across UC health centers (more details at https://www.ucop.edu/uc-health/departments/center-for-data-driven-insights-and-innovations-cdi2.html or by contacting).

## Acknowledgements

The authors thank the Center for Data-driven Insights and Innovation at UC Health (CDI2; https://www.ucop.edu/uc-health/functions/center-for-data-driven-insights-and-innovationscdi2.html), for its analytical and technical support related to use of the UC Health Data Warehouse and related data assets, as well as Innovation Clinical and Translational Science Institutes of UCSF. They thank Dr. Michael Kattah for supporting the review of manuscript.

## Funding

No external funding was received for conducting this study

## Author Contributions

All authors participated in the study design. Danny Hoi Tsun Chu performed data analysis. Danny Hoi Tsun Chu and Ann-Kathrin Schalkamp contributed to drafting the manuscript and figures. All authors revised and approved the final version of the manuscript. Vivek A. Rudrapatna supervised the research.

## Competing interest declaration

Vivek A. Rudrapatna previously received funding from BeOne Medicines Ltd, Janssen, Takeda, Blueprint Medicines, Merck, Mitsubishi Tanabe, Genentech, and Stryker. He is involved with ZebraMD, AucucareAI, and Data Unite. He has previously received payment from Natera and Ironwood. The remaining authors disclose no conflicts. The authors declare that no actual competing interests exist.

## Data availability

Access to UCDDP and UCSF CDW database are controlled due to sensitive nature of the patient data, and they were protected by law. The UCSF CDW database can be accessed by UCSF-affiliated individuals by contacting UCSF Clinical and Translational Science Insitute or UCSF’s Information Commons team, more information could be found at https://data.ucsf.edu/research/ucsf-data. If the reader is unaffiliated with UCSF, they can set up an official collaboration with a UCSF-affiliated investigator by contacting the principal investigator.

UCDDP is only available to UC researchers who have completed analyses in their respective UC first and have provided justification for scaling their analyses across UC health centers (more details at https://www.ucop.edu/uc-health/departments/center-for-data-driven-insights-and-innovations-cdi2.html or by contacting).

## Ethical Statement

This study complies with all relevant ethical regulations, and was approved by Institutional Review Board of University of California, San Francisco, with approval reference #18-24588.

## Notes

### Funding Statement

This study did not receive any funding

