## Supplementary material for "Risk factors for immune checkpoint inhibitor colitis: a retrospective multi-center cohort study using electronic health records": All supplemental files

**Supplemental table 1. ICD code used**

| **Diagnosis** | **ICD-9 code** | **ICD-10 code** |
| --- | --- | --- |
| GI-related tumors | 150 – 159 | C15 – C26 |
| Pregnancy | 630 – 679 | Z32 – Z39, Z3A |
| Immune disorder | 576.1, 686.01, 571.42, 579.0, 720.0, 446.4, 571.6, 379, 364.0, 535.1, 696.1, 446.0, 714.0, 135, 493, 446.5, 696.0, 242.0, 725, or contain the diagnosis with the name of Primary sclerosing cholangitis, Pyoderma gangrenosum, Autoimmune hepatitis, Celiac disease, Ankylosing spondylitis, Churg Strauss syndrome, Primary biliary cholangitis, Episcleritis, Iridocyclitis, Atrophic gastritis, Psoriasis, Polyarteritis nodosa, Rheumatoid arthritis, Type 1 diabetes, Sarcoidosis, Asthma, Giant cell arteritis, Psoriatic arthritis, Grave's disease, Polymyalgia rheumatica | K83.01, L88, K75.4, K90.0, M45, M30.1, K74.3, H15.1, H20.0, K29.4, L40, M30.0, M06.9, E10, D86, J45, M31.6, L40.5, M07, E05.0, M35.3, or contain the diagnosis with the name of Primary sclerosing cholangitis, Pyoderma gangrenosum, Autoimmune hepatitis, Celiac disease, Ankylosing spondylitis, Churg Strauss syndrome, Primary biliary cholangitis, Episcleritis, Iridocyclitis, Atrophic gastritis, Psoriasis, Polyarteritis nodosa, Rheumatoid arthritis, Type 1 diabetes, Sarcoidosis, Asthma, Giant cell arteritis, Psoriatic arthritis, Grave's disease, Polymyalgia rheumatica |
| Gastroesophageal reflux disorder | 530.81 | K21 |
| Gastrointestinal ulcer | 531 – 534 | K25 – K28 |
| Irritable Bowel Syndrome | 564.1 | K58 |
| Liver disease | 570 - 573 | K70 - K79 |
| Pancreas, gallbladder or other biliary tract disease | 574 - 577 | K80 - K89 |
| Other gastrointestinal disorder | Anything starts with 530-579 but not stated above or inflammatory bowel disease | Anything starts with K20-99 but not stated above or inflammatory bowel disease |
| Melanoma | 172 or contain the diagnosis with the name melanoma | C43 or contain the diagnosis with the name melanoma |
| Other skin cancer | 173 | C44 |
| Non-small cell lung cancer | 162 with non-small cell lung cancer/NSCLC stated in the diagnosis name | C34 with non-small cell lung cancer/NSCLC stated in the diagnosis name |
| Small cell lung cancer | 162 with small cell lung cancer stated in the diagnosis name | C34 with small cell lung cancer stated in the diagnosis name |
| Renal cell carcinoma | 189 | C64 |
| Head and neck cancer | 140-148, 160, 161, 162.0, 195.0 or 195.1 (Any diagnosed tumor of the lip, oral cavity, pharynx, nasal cavities, larynx, and other related regions in the head, face, neck and thorax) | C00 – 13, C30 – 33, C76.0 or C76.1 (Any diagnosed tumor of the lip, oral cavity, pharynx, nasal cavities, larynx, and other related regions in the head, face, neck and thorax) |
| Depression | 296.2, 296.3, 300.4, 311 | F32, F33, F34.1, F41.2 |
| Bipolar disorder | 296.0, 296.4 – 296.8 | F31 |
| Anxiety | 300.00 - 02, 300.22 – 23, 300.29 | F41 |
| Other mood disorder | 296 but not 296.0, 296.2 - 296.8 | F30, F34 – 39 (except 34.1) |
| Schizophrenia and related psychosis | 295, 297, 298 (Any diagnosis of schizophrenia, delusional disorders, and other non-mood psychotic disorders) | F20 – F29 (Any diagnosis of schizophrenia, delusional disorders, and other non-mood psychotic disorders) |

Clinical characteristics of UCSF CDW cohort

We identified 5,301 patients who received checkpoint inhibitors, of which 2,778 had one of our defined cancers and no history of GI-related tumors. We further excluded 327 patients who were younger than 18 years before their first administration of CI and 156 with insufficient data (no date). Our final cohort included 2,167 patients (Fig. 1), 39.1% female, median age of 67.40 (IQR: 58.2 – 75.4), with a median follow-up of 8 months (<1-108 months) (Table 3). 222 (10.24%) developed colitis during follow-up (Fig. 2). The cumulative incidence of colitis following CI administration was 10.6% (95% CI: 9.07 to 12.18%) after 12 months, given that competing events do not happen. The median time to colitis onset was 5 months (IQR: 2.25 – 10.75 months).
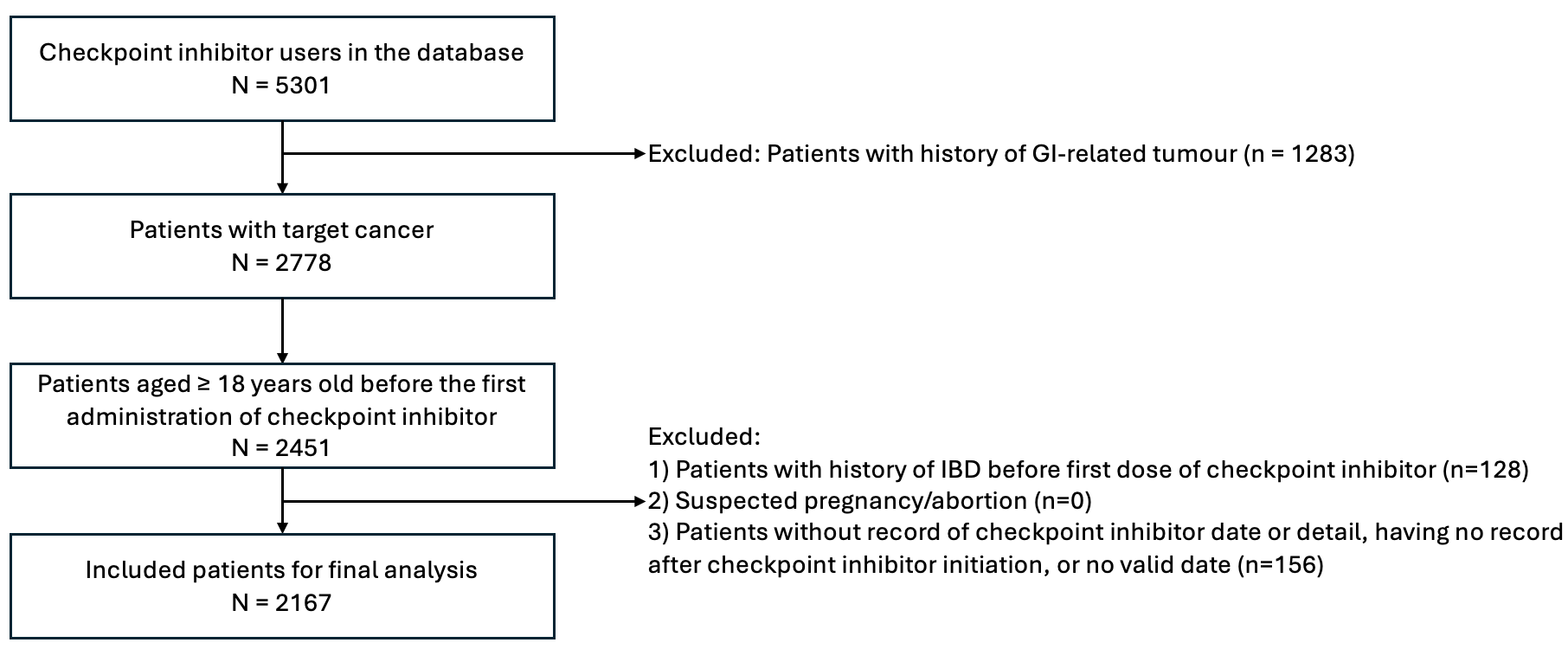

**Supplemental fig 1. Flow chart of patient selection and data extraction**

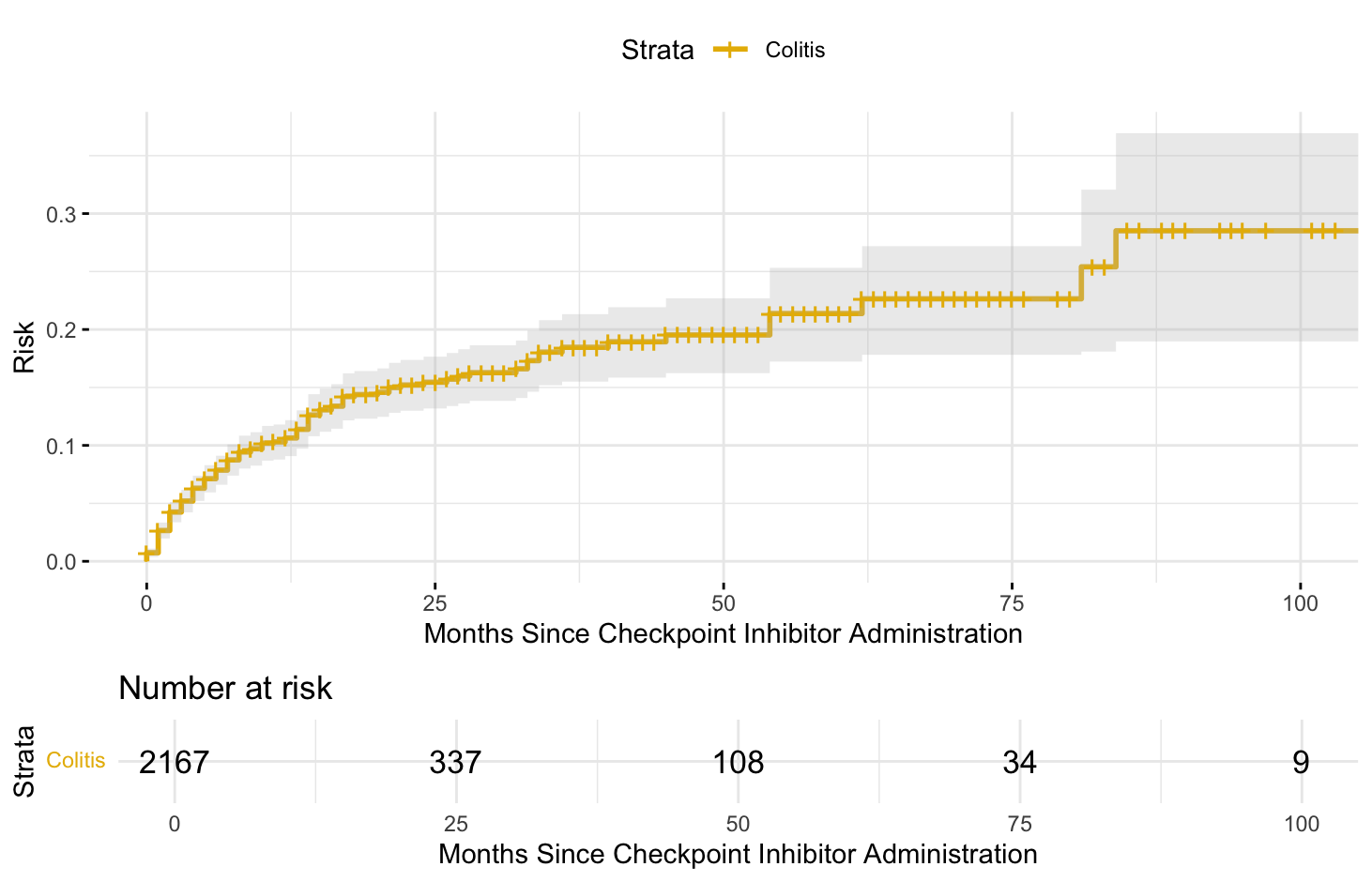

**Supplemental fig 2. Cumulative incidence curve of colitis of the UCSF CDW cohort**

**Supplemental table 2. Missingness of the dataset that requires imputation in UCSF CDW**

For each variable where data was missing, the number of missing entries and the percentage are shown.

| Predictor | Number missing (%) |
| --- | --- |
| Race/Ethnicity | 605 (1.45%) |
| Distance / Driving time to San Francisco General Hospital / Area Deprivation Index State Rank | 1033 (2.47%) |
| Smoking status | 76 (0.18%) |
| Marital status | 785 (1.88%) |
| BMI before ICI administration | 640 (1.53%) |
| BMI (time-varying) | 14615 (34.8%) (before imputing with linear interpolation)  194 (0.46%) (after imputing with linear interpolation) |

**Supplemental table 3. Participant characteristics for University of California, San Francisco cohort extracted from Central Data Warehouse**

The demographics and characteristics of the identified UCSF CDW cohort are shown.

| **Variable** | **Overall** | **Non-CIC** | **CIC** |
| --- | --- | --- | --- |
| N | 2167 | 1945 | 222 |
| Race: White(%) | 1484 (68.5) | 1315 (67.6) | 169 (76.1) |
| Sex: Female(%) | 847 (39.1) | 748 (38.5) | 99 (44.6) |
| Age (median (IQR)) | 67.40 (58.20 - 75.40) | 67.60 (58.40 - 75.50) | 65.60 (56.20 - 74.08) |
| Weighted mean driving hours to SF in each 3-digit zip code (Median (IQR))* | 0.94 (0.60 - 1.62) | 0.94 (0.60 - 1.62) | 0.94 (0.64 - 1.78) |
| Weighted mean miles to SF in each 3-digit zip code (Median (IQR))* | 38.70 (17.50 - 79.62) | 38.70 (17.50 - 79.62) | 38.70 (22.70 - 92.95) |
| Weighted median ADI State Rank in each 3-digit zip code (Median (IQR)) | 10.00 (2.00 - 12.00) | 10.00 (2.00 - 12.00) | 10.00 (2.00 - 16.50) |
| Smoking Status (%) |  |  |  |
| Never smoked | 988 (45.6) | 878 (45.1) | 110 (49.5) |
| Former smoker | 1068 (49.3) | 963 (49.5) | 105 (47.3) |
| Current smoker | 111 (5.1) | 104 (5.3) | 7 (3.2) |
| Marital Status (%) |  |  |  |
| Partnered | 1408 (65.0) | 1259 (64.7) | 149 (67.1) |
| Separated/Divorced | 198 (9.1) | 168 (8.6) | 30 (13.5) |
| Widowed | 202 (9.3) | 187 (9.6) | 15 (6.8) |
| Single | 359 (16.6) | 331 (17.0) | 28 (12.6) |
| BMI before CI administration (Mean(SD)) | 26.43 (5.78) | 26.38 (5.72) | 26.81 (6.26) |
| ICI regimen ever used (%) |  |  |  |
| Anti-PD-1/L1 only | 2005 (92.5) | 1827 (93.9) | 178 (80.2) |
| Anti-CTLA-4 only | 26 (1.2) | 19 (1.0) | 7 (3.2) |
| Combined | 391 (18.0) | 313 (16.1) | 78 (35.1) |
| Last ICI regimen used (%) |  |  |  |
| Anti-PD-1/L1 only | 1754 (80.9) | 1617 (83.1) | 137 (61.7) |
| Anti-CTLA-4 only | 22 (1.0) | 15 (0.8) | 7 (3.2) |
| Combined | 391 (18.0) | 313 (16.1) | 78 (35.1) |

Coefficients

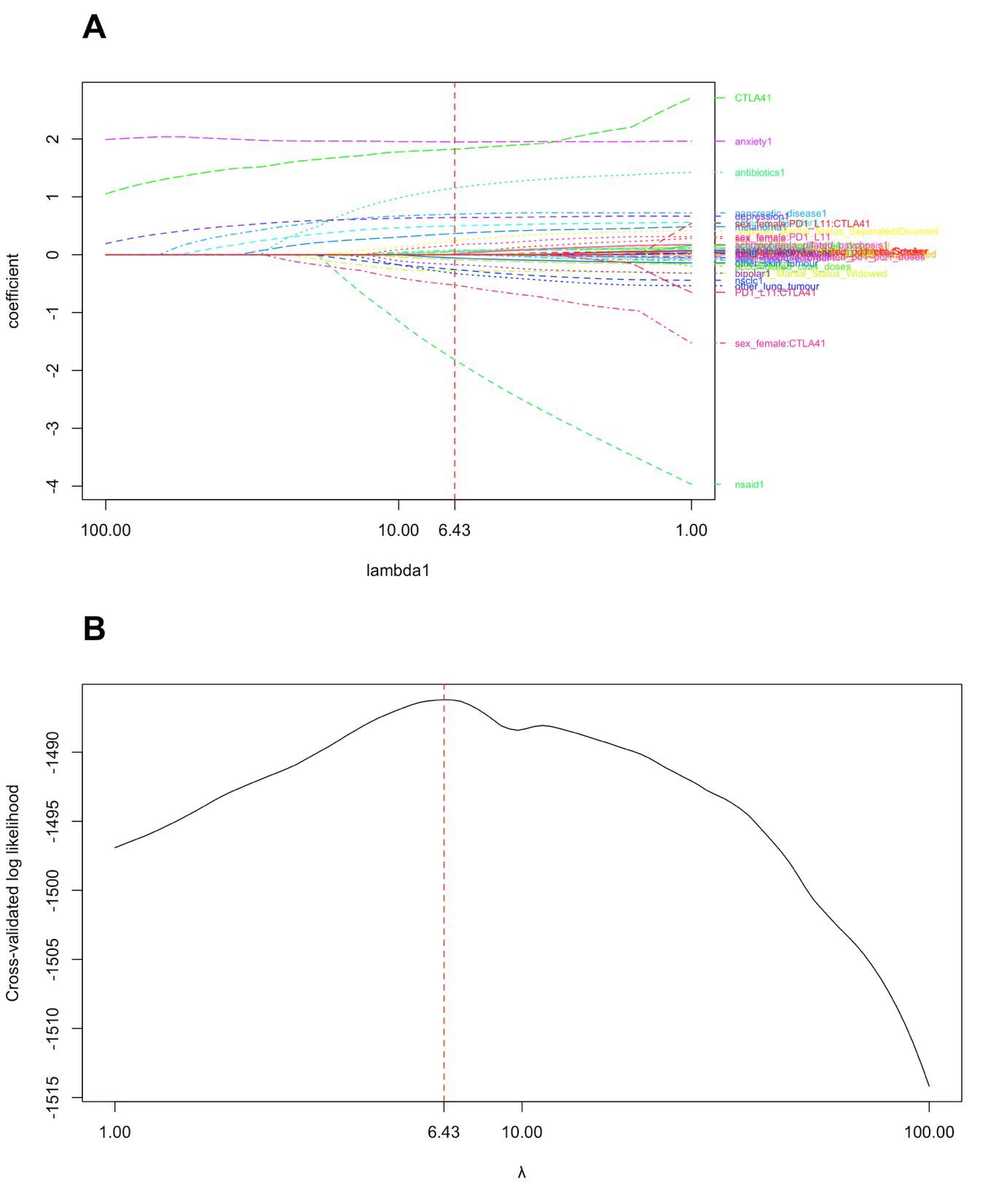

**Supplemental fig 3. A) Coefficient Estimates Across Explored Lambda Values in Lasso Cox Regression B) Change of cross-validated log-likelihood along lambda values**

The figures indicated the selection of hyperparameter, λ, for LASSO and the change of coefficient and the 10-fold cross-validated likelihood.

**Supplemental table 4. LASSO Cox regression coefficient from the UCSF CDW Dataset**

The table denoted the shrunk coefficient after applying LASSO Cox regression to the UCSF CDW dataset.

| **Feature** | **Coefficient** |
| --- | --- |
| Race: White | 0 |
| Sex: female | 0 |
| Smoking Status |  |
| Current smoker | 0 |
| Former smoker | 0 |
| Non-smoker | (ref) |
| Age | 0 |
| Mean driving miles of a 3-digit zip code to UCSF | -0.000736 |
| Mean driving hours of a 3-digit zip code to UCSF | 0 |
| Median ADI State Rank of a 3-digit zip code | 0.009252 |
| Marital Status |  |
| Partnered | 0.051918 |
| Separated/Divorced | 0.240732 |
| Widowed | -0.274043 |
| Single | (ref) |
| BMI (baseline) | 0.057377 |
| BMI (time-varying) | -0.085129 |
| Anti-PD-1/L1 | 0 |
| Anti-CTLA-4 | 1.823714 |
| Anti-PD-1/L1 x Anti-CTLA-4 | 0 |
| Accumulated number of Anti-PD-1/L1 administration | 0.007235 |
| Accumulated number of Anti-CTLA-4 administration | 0 |
| Accumulated number of combined therapy administration | 0 |
| NSAID use | -1.814662 |
| Antibiotics use | 1.153141 |
| Immune disorder | 0.076288 |
| Gastroesophageal Reflux Disorder | 0.048444 |
| Ulcer | 0 |
| Irritable Bowel Syndrome | 0 |
| Liver Disease | 0 |
| Disorder of gallbladder, other biliary tract, or pancreas | 0.701032 |
| Other GI disorder | 0.497537 |
| Melanoma | 0.366219 |
| Other skin tumour | -0.055516 |
| Non-small cell lung cancer | -0.256228 |
| Small cell lung cancer | -0.322122 |
| Renal Cell Carcinoma | 0 |
| Head and neck cancer | 0 |
| Appendectomy | 0 |
| Depression | 0.641844 |
| Bipolar disorder | -0.169765 |
| Other mood disorder | 0 |
| Anxiety | 1.950846 |
| Schizophrenia and related psychosis | 0.012234 |
| Female x Anti-PD-1/L1 | 0.162006 |
| Female x Anti-CTLA-4 | -0.525229 |
| Female x Anti-PD1/L1 x Anti-CTLA-4 | 0 |
| Female x Accumulated Anti-PD-1/L1 administration | 0 |
| Female x Accumulated Anti-CTLA-4 administration | 0 |
| Female x Accumulated combined therapy administration | 0 |

**Supplemental table 5. Incidence Rate of the UCDDP dataset for each site**

The table recorded the incidence rate from each site under each institution in the UC system

| Site | Incidence Rate (95% confidence interval) per 100 person-year |
| --- | --- |
| Site 1 | 11.60 (9.70 – 13.75) |
| Site 2 | 8.89 (7.46 – 10.52) |
| Site 3 | 8.10 (7.21 – 9.07) |
| Site 4 | 46.2 (1.17 – 257.15) |
| Site 5 | 14.2 (12.56 – 15.96) |
| Site 6 | 8.83 (7.86 – 9.89) |

**Supplemental table 6. Missingness of the dataset that requires imputation in UCDDP**

The table below showed the missing information of UCDDP. For each variable where data was missing, the number of missing entries and the percentage is shown.

| **Predictor** | **Number missing (%)** |
| --- | --- |
| Race/Ethnicity | 0 (0%) |
| Marital status | 223 (0.16%) |
| BMI before ICI administration | 49 (0.03%) |
| BMI (time-varying) | 15198 (10.5%) (before imputing with linear interpolation)  1132 (0.78%) (after imputing with linear interpolation) |

**Supplemental table 7: Coefficients and hazard ratios of the Cox regression model before and after adding institution as predictors**

The table below shows the difference between each covariates with regards to the coefficient and hazard ratio

| **Covariates** | **Cox without institution coefficient** | **Cox with institution coefficient** | **Cox without institution hazard ratio** | **Cox with institution hazard ratio** | **Percentage change in hazard ratio** |
| --- | --- | --- | --- | --- | --- |
| Anti-CTLA4 | 1.299059 | 1.268268 | 3.67 | 3.55 | -3.03% |
| Accumulated PD1/L1 dose | 0.009084 | 0.009563 | 1.01 | 1.01 | 0.05% |
| Antibiotics | 1.000813 | 1.006738 | 2.72 | 2.74 | 0.59% |
| NSAIDs | 0.405117 | 0.402546 | 1.50 | 1.50 | -0.26% |
| GORD | 0.269947 | 0.271742 | 1.31 | 1.31 | 0.18% |
| Immune disorder | 0.044848 | 0.037491 | 1.05 | 1.04 | -0.73% |
| Other GI | 0.276693 | 0.266615 | 1.32 | 1.31 | -1.00% |
| IBS | 0.285535 | 0.287438 | 1.33 | 1.33 | 0.19% |
| Depression | 0.18601 | 0.175051 | 1.20 | 1.19 | -1.09% |
| Bipolar Disorder | -0.339231 | -0.335415 | 0.71 | 0.72 | 0.38% |
| Anxiety | 0.285882 | 0.267002 | 1.33 | 1.31 | -1.87% |
| Schizophrenia and related psychosis | -0.112869 | -0.131483 | 0.89 | 0.88 | -1.84% |
| Melanoma | 0.161015 | 0.187306 | 1.17 | 1.21 | 2.66% |
| Other skin cancer | -0.097886 | -0.106605 | 0.91 | 0.90 | -0.87% |
| NSCLC | -0.069457 | -0.100101 | 0.93 | 0.90 | -3.02% |
| SCLC | -0.223606 | -0.212467 | 0.80 | 0.81 | 1.12% |
| BMI (baseline) | 0.056053 | 0.054793 | 1.06 | 1.06 | -0.13% |
| BMI (time-varying) | -0.074302 | -0.07437 | 0.93 | 0.93 | -0.01% |
| Partnered | 0.08192 | 0.082279 | 1.09 | 1.09 | 0.04% |
| Separated | 0.052915 | 0.044705 | 1.05 | 1.05 | -0.82% |
| Widowed | -0.098812 | -0.103883 | 0.91 | 0.90 | -0.51% |
| Single | (ref) | (ref) | (ref) | (ref) | NA |
| Site 1 | NA | -0.168313 | NA | 0.85 | NA |
| Site 2 | NA | -0.20907 | NA | 0.81 | NA |
| Site 3 | NA | 0.171853 | NA | 1.19 | NA |
| Site 4 | NA | 0.108479 | NA | 1.11 | NA |
| Site 5 | NA | 1.463878 | NA | 4.32 | NA |
| Site 6 | (ref) | (ref) | (ref) | (ref) | NA |
| CTLA4 x female | -0.143848 | -0.152359 | 0.87 | 0.86 | -0.85% |
| PD1/L1 x female | 0.184691 | 0.189838 | 1.20 | 1.21 | 0.52% |

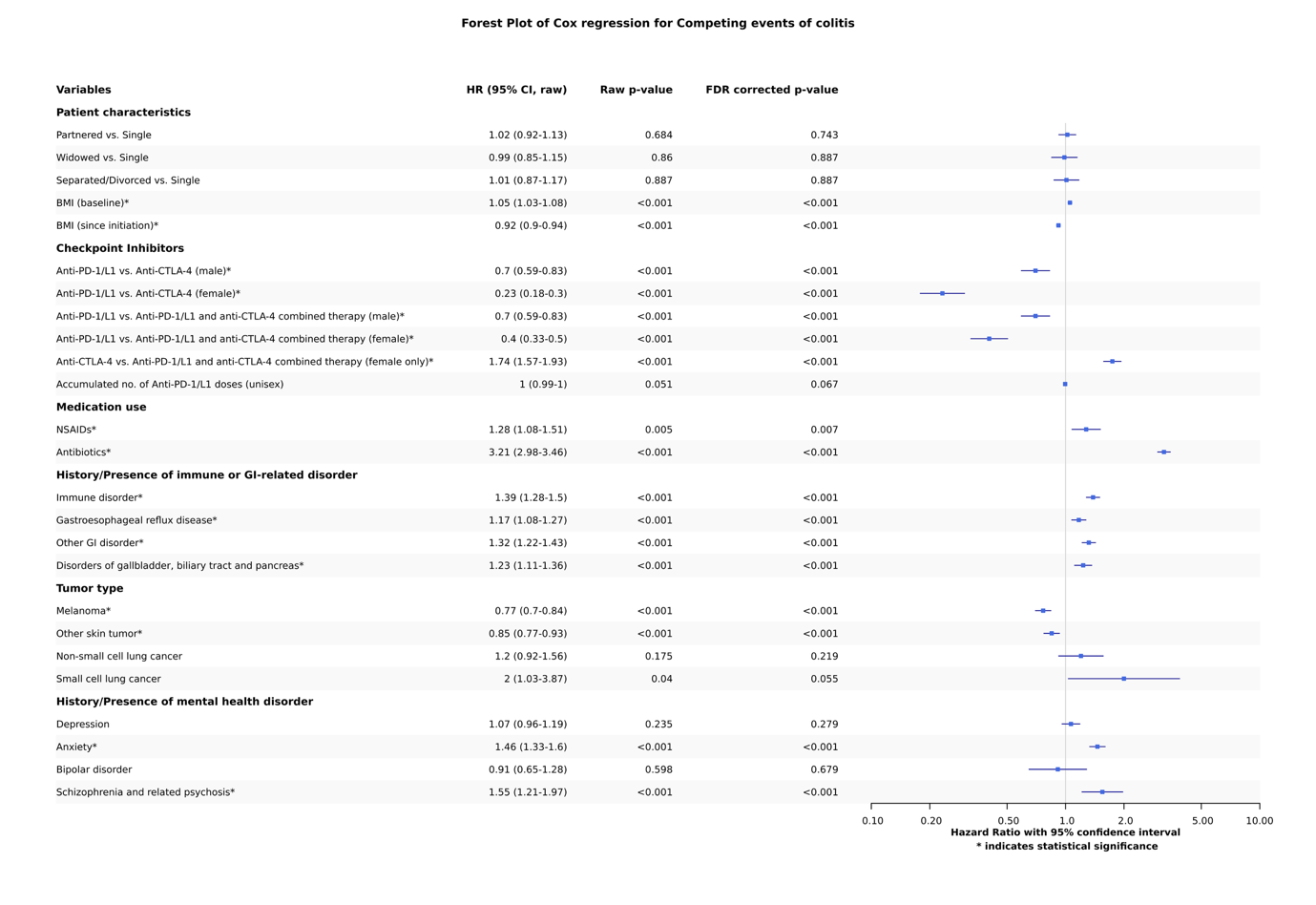

**Supplemental fig. 4 Forest plot of the Cox regression model applied for competing event**

The estimated coefficients of the Cox regression model are shown with their Hazard ratio (HR) and respective 95% Confidence interval as well as their uncorrected and FDR-corrected p-value.

**Supplemental table 8: Post-hoc analysis for diagnoses under Other GI disorders**

The table showed, from the most frequent to least frequent colitis case happened for each diagnosis under “other GI disorder”

| **Diagnoses** | **ICD-10 code** | **Colitis count** | **Total count** | **Frequency (%)** |
| --- | --- | --- | --- | --- |
| Vascular disorder of intestine | K55 | <10 | 18 | 33.33 |
| Abscess/Perforation/Fistula/Ulcer of intestine | K63.1 – 63.4 | 11 | 43 | 25.58 |
| Dysplasia of anus | K62.82 | <10 | 17 | 23.53 |
| Other specified functional intestinal disorders | K59.8 | <10 | 16 | 18.75 |
| Vomiting | R11 | <10 | 47 | 17.02 |
| Appendix disorder | K35 – 38 | <10 | 54 | 16.67 |
| Other specificed disorders of peritoneum | K66.8 | 17 | 103 | 16.5 |
| Peritonitis | K65 | 12 | 77 | 15.58 |
| Rectal prolapse | K62.3, | <10 | 13 | 15.38 |
| Peritoneal adhesion (postprocedural/postinfection) | K66.0 | 22 | 145 | 15.17 |
| Hemorrhage of anus and rectum | K62.5 | 26 | 172 | 15.12 |
| Ileus and intestinal obstruction without hernia | K56 | 63 | 425 | 14.82 |
| Other specified diseases of the digestive system | K92.8 | 97 | 676 | 14.35 |
| Neurogenic bowel | K59.2 | <10 | 29 | 13.79 |
| Hemorrhoids and perianal venous thrombosis | K64 | 91 | 660 | 13.79 |
| Functional diarrhea | K59.1 | <10 | 22 | 13.64 |
| Diverticular disease of intestine | K57 | 210 | 1547 | 13.57 |
| Other diseases of stomach and duodenum | K31 | 41 | 313 | 13.1 |
| Rectal polyp | K62.1 | 10 | 77 | 12.99 |
| Polyp of colon | K63.5 | 40 | 311 | 12.86 |
| Other specified diseases of anus and rectum | K62.8 | 20 | 159 | 12.58 |
| Abscess of anal and rectal regions | K61 | <10 | 56 | 12.5 |
| Gastritis and duodenitis | K29 | 44 | 353 | 12.46 |
| Melena | K92.1 | 31 | 249 | 12.45 |
| Functional dyspepsia | K30 | 28 | 232 | 12.07 |
| Hernia | K40 | 194 | 1623 | 11.95 |
| Intestinal malabsorption | K90 | <10 | 42 | 11.9 |
| Intraoperative and postprocedural complications and disorders of digestive system, not elsewhere classified | K91 | 116 | 995 | 11.66 |
| Constipation | K59.0 | 413 | 3699 | 11.17 |
| Esophageal disorder | K20, K22, K23 | 75 | 694 | 10.81 |
| Gastrointestinal hemorrhage, unspecified | K92.2 | 18 | 171 | 10.53 |
| Hematemesis | K92.0 | <10 | 63 | 9.52 |
| Complications of artificial openings of the digestive system | K94 | 26 | 311 | 8.36 |
| Disorders of retroperitoneum | K68 | <10 | 36 | 8.33 |
| Complications of bariatric procedures | K95 | <10 | 84 | 8.33 |
| Hemoperitoneum | K66.1 | <10 | 25 | 8 |
| Anal or rectal pain/Anal spasm | K62, K59.4 | <10 | 63 | 7.94 |
| Fissure and fistula of anal and rectal regions | K60 | <10 | 29 | 3.45 |
